# Symptom burden, viral load, and antibody response to ancestral SARS-CoV-2 strain [D614G] in an outpatient household cohort

**DOI:** 10.1101/2024.10.27.24316219

**Authors:** Mehal Churiwal, Kathleen Tompkins, Gabrielle Streeter, Christy Litel, Sydney Mason, Kelly Lin, Meredith Muller, Srijana Chhetri, Tia Belvin, Christopher Basham, Maureen Whittelsey, Tyler Rapp, Lakshmanane Premkumar, Carla Cerami, Jessica T. Lin

**Affiliations:** Institute of Global Health and Infectious Diseases, University of North Carolina School of Medicine, Chapel Hill, NC USA; Department of Microbiology and Immunology, University of North Carolina School of Medicine, Chapel Hill, NC USA; Medical Research Council Unit The Gambia at the London School of Hygiene & Tropical Medicine, The Gambia

## Abstract

**Background:** Early in the SARS-CoV-2 pandemic, description of COVID-19 illness among non-hospitalized patients was limited. Data from household cohorts can help reveal the full spectrum of disease and the potential for long-term sequelae, even in non-severe disease.

**Methods:** Daily symptom diaries were collected in a US household cohort of SARS-CoV-2 infection from April to November 2020, during the pre-COVID vaccine period. SARS-CoV-2 nasal viral loads were measured at study entry and weekly until day 21; serologic testing was performed at study entry and day 28. A subset of volunteers underwent an additional assessment 8-10 months later. Participants who met the criteria for early infection—testing antibody-negative at study entry but PCR-positive either at baseline or during follow-up—were included in this analysis (n=143).

**Results:** Daily symptoms were ascertained in 143 outpatients with acute COVID-19, including 60 index cases who sought testing and 83 of their household contacts. Asymptomatic cases comprised 16% (13/83) of SARS-CoV-2 infections detected among household contacts. Among 119 persons with mild or moderate illness, the number of symptoms peaked 3 or 4 days after symptom onset. Fever and anosmia occurred in nearly half of participants. Symptom severity was associated with increased age, viral load, and cardiovascular disease. Increased BMI was associated with a higher antibody level at day 28, independent of symptom severity. Those with a higher day 28 antibody level were more likely to develop symptoms consistent with post-acute sequelae of SARS-CoV-2 (PASC), also known as long COVID-19, 8-10 months later.

**Conclusions:** Fever, anosmia, as well as asymptomatic infection were common features of COVID-19 non-severe illness when the D614G variant circulated in the US, before the availability of vaccines or outpatient therapies. Antibody levels following acute infection were linked to the development of symptoms of PASC 8-10 months later.

## Introduction

At the onset of the COVID-19 pandemic in 2019-2020, research on its clinical course, viral load, and antibody responses to SARS-CoV-2 largely focused on patients who presented to or were admitted to hospitals and medical centers [1–3]. Since an estimated 81% of infections caused no or mild pneumonia [4], these descriptions were limited in their ability to describe the full spectrum of disease. Detailed descriptions of symptom spectrum and severity are needed to best understand whether clinical patterns of disease are changing as the virus evolves and new variants emerge. They are also useful for determining who may be at risk for post-acute sequelae of SARS-CoV-2 infection (PASC) given the large number of persons contracting non-severe disease throughout the pandemic.

Household cohorts, in which family members (household contacts) are followed after an index case is diagnosed with SARS-CoV-2 infection, provide the ideal study design for capturing acute infection in secondary cases that better reflect the full spectrum of disease. The UNC CO-HOST Study (COVID-19 Household Transmission Study) was an observational cohort study involving approximately 100 households across a diverse socio-demographic cohort in central North Carolina with the primary aim of determining secondary household attack rates in households where infected individuals self-isolated at home [5]. Here, we report the results from individuals with SARS-CoV-2 infection and their PCR-positive household contacts who tested positive over the course of the study. We describe the course of symptoms, quantitative nasal viral loads, and antibody responses over a 28-day period. We additionally evaluated vaccination status, post-acute sequelae, and antibody titers in a subset of participants 8-10 months after acute infection.

## Methods

### Study design and sample collection

Details of the UNC CO-HOST study have been described previously [5]. Briefly, between 28/04/2020 and 20/11/2020, participants were enrolled in a prospective longitudinal cohort study. Index cases were enrolled through a COVID-19 testing center at the University of North Carolina at Chapel Hill that provided PCR results within 24 hours, allowing for timely study enrollment. Index cases were eligible for enrollment if they were at least 18 years of age, had a positive COVID-19 PCR test at the testing center, and lived within a 1-hour driving distance of the university. Household contacts at least 1 year of age living in the same home as the index patient were invited to enroll if they intended to remain in the home for the 28-day study period. Pre-screening was conducted via phone, and enrollment and study visits were conducted outside participants’ homes using a mobile medical van.

Incident SARS-CoV-2 infection in the household was detected based on PCR testing at baseline and weekly PCR thereafter until study day 21, as well as serologic testing at study entry and 28 days later. At study entry, all study participants received nasopharyngeal (NP) and nasal mid-turbinate (NMT) swabs for PCR testing, with instructions on how to self-perform NMT swabs for future visits. Quantitative PCR testing was performed using a CDC RT-qPCR protocol authorized for emergency use [6]. NP viral loads were mathematically transformed into corresponding NMT viral load values based on data from paired samples [7]. At study entry, participants also had venipuncture for serology testing by enzyme-linked Immunosorbent Assay (ELISA) and a finger stick for rapid point-of-care lateral flow assay antibody testing (Biomedomics, Morrisville, NC). ELISA testing was performed using an in-house assay targeting the SARS-CoV-2 receptor binding domain [8]. On days 7, 14, and 21 of the study, participants performed self-collected NMT swabs for viral load PCR testing. On day 28 of the study, participants underwent repeat blood collection for serology by ELISA. All samples were immediately placed on ice and transferred to a BSL-2 laboratory for processing within 2 hours.

### Symptom evaluation

Participants completed a baseline questionnaire that included an assessment of symptoms, and they subsequently completed electronic daily symptom diaries that were sent via email. In the baseline questionnaire, participants were asked to quantify the number of days symptoms were present for any symptom reported. Symptom diaries consisted of a checklist of 14 symptoms in both English and Spanish that the participant completed if they answered “Yes” to the question “Are you having any symptoms?” These 14 symptoms were: fever, chills, muscle aches, runny nose, sore throat, loss of taste or smell, cough, shortness of breath, chest pain, wheezing, diarrhea, vomiting/nausea, headache, and abdominal pain.

Index cases completed daily symptom diaries until no symptoms were reported for two consecutive days. Household contacts received daily symptom evaluations until study day 21 to evaluate for symptoms that could be consistent with new infection. If a participant failed to fill out a symptom diary for two consecutive days, they were contacted by study staff to facilitate completion. Symptom evaluation was performed over the phone for participants who indicated this preference. Questionnaires and symptom diaries for minors were sent to and completed by their designated guardians if no separate email for the minor was provided. If a participant was hospitalized, they remained in the study for outcome assessment, but no further samples or questionnaires were collected/sent.

### Extended follow up

Participants were re-contacted between 8-10 months after their initial enrollment, between 05/05/2021 and 27/08/2021, for a voluntary follow-up assessment. This consisted of a single visit during which they completed a questionnaire on their current state of health, persistence of 10 symptoms potentially attributable to post-acute sequelae of SARS-CoV-2 infection (PASC or “long COVID”), additional documented COVID-19 infections, and receipt of the COVID-19 vaccine. Additionally, they reported their current state of health, additional documented COVID-19 infections, and receipt of the COVID-19 vaccine. They also had venipuncture for ELISA antibody testing.

### Ethics, standards, and informed consent

The study was approved by the Institutional Review Board at the University of North Carolina and is registered at clinicaltrials.gov (NCT04445233). All participants (or their parents/guardians) gave written, informed consent. Minors over the age of 7 provided assent.

### Statistical analysis

The results presented here detail the reported symptoms, viral loads, and antibody responses of participants who tested positive for SARS-CoV-2 immediately prior to study entry or during the 28-day duration of the study. We considered individuals to be recently or newly infected if they had a positive viral load on any nasal swab and were antibody-negative at study entry, or if they seroconverted during the course of the study. Individuals who were seropositive by either ELISA or rapid antibody testing at study entry were excluded from analysis as they may represent prior infections or late enrollment of a primary infection and thus may have been past their symptom and viral load peaks.

Data were collected into a RedCap database (Vanderbilt University) and analyzed using STATA software (College Station, TX) and R 4.0.2 (R Core Team, Vienna, Austria). Day of symptom onset in relation to study entry was based on either a survey administered by the UNC diagnostic center regarding symptom duration for each symptom endorsed or a similar question in the baseline questionnaire (whichever was earliest); for incident cases in the household, day of symptom onset was captured in real-time through the daily symptom diaries.

Participants were categorized as being asymptomatic, or having mild, moderate, or severe/hospitalized symptoms based on a modified CDC definition [9]. Symptoms were quantified cumulatively throughout the study to capture both the number of symptoms per day and the duration of symptoms. For example, if an individual reported 1 symptom on day 1, 3 symptoms on day 2, and 1 symptom on day 3, they would be considered to have 5 cumulative symptoms. Participants were considered asymptomatic if they reported at most 1 cumulative symptom for the duration of their enrollment. Additionally, the symptom reported could not be anosmia, dyspnea, or new or worsening cough. Participants were considered to have mild illness if they reported between 2 and 10 cumulative symptoms or 1 symptom of anosmia, dyspnea, or new or worsening cough. Participants were considered to have moderate illness if they reported 11 or more cumulative symptoms but were not hospitalized and severe illness if they were hospitalized or died during the study. Since symptom diaries were not collected once a participant was hospitalized, 10 symptoms for each day of hospitalization were imputed for study analyses. The number of cumulative symptoms on any given day starting from the day of onset was plotted to depict the pattern of illness in those with different disease severity. Calculations were only done on true reported data. On days when symptom diaries were not filled out, the data was treated as null, and the other complete symptom diaries were used to inform calculations. Overall, there was <10% missing data from symptom diaries.

We examined the frequency and duration of twelve commonly reported symptoms. Associations between symptom severity and participant characteristics were calculated using one-way ANOVA or chi-squared testing. Associations between viral load (highest viral load measured, usually on day of enrollment for index cases), antibody responses, and patient characteristics were calculated using simple linear regression or one-way ANOVA. Associations between the presence of PASC symptoms, antibody status, and patient characteristics were assessed using Mann-Whitney U or chi-squared testing. All COVID-positive individuals from the original household cohort that were followed up at 8-10 months were included in the PASC analysis, regardless of whether they were included in the analysis of those with acute infection.

## Results

### Symptom severity

Between April 28 and November 20, 2020, 315 individuals from 102 households were enrolled in the UNC CO-HOST Study. Of these, 143 individuals met our criteria for early SARS-CoV-2 infection (PCR-positive and antibody-negative at study entry) and had no more than two days of missing symptom diaries. This included both 60 index cases who sought testing and 83 of their household contacts, among which 13/143 (9%) were asymptomatic, 53/143 (37%) had mild illness, 68/143 (48%) had moderate disease, and 9/143 (6%) were hospitalized (Figure 1). Of the nine who were hospitalized, two ultimately died. Asymptomatic infections comprised 16% (13/83) of household contacts diagnosed by active surveillance. Secondary infections detected in household contacts had fewer cumulative symptoms than index cases who presented to care (cumulative 9.7 vs. 16.4 symptoms, respectively, p<0.001). Those with mild or moderate illness reported the most symptoms 3 or 4 days after symptom onset, while those with severe disease or who were hospitalized experienced escalating symptoms over more than one week (Fig 1).

**Fig 1.**
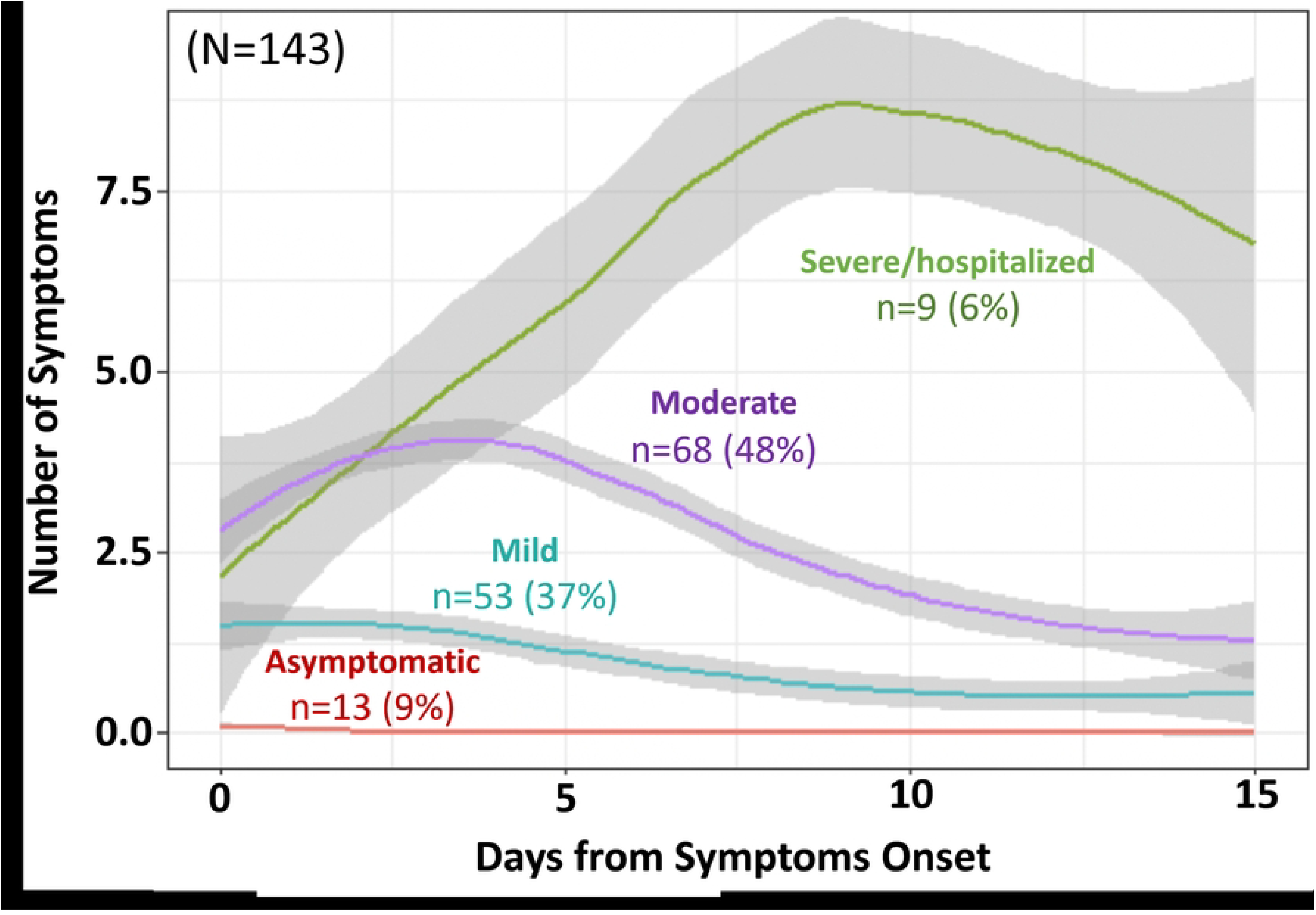
COVID-19 symptom progression by severity of illness. 143 study participants that met criteria for newly positive SARS-CoV-2 infection (PCR positive and antibody-negative at study entry) were categorized into symptom severity categories based on the number of cumulative symptoms reported in daily symptom diaries. For the asymptomatic category, days from study enrollment was substituted for days from symptom onset.

### Frequency, onset, and duration of symptoms

Participants reported a wide array of symptoms attributable to COVID-19 illness, with more nonspecific symptoms such as cough, headache, and myalgias early in illness and dyspnea and anosmia presenting later in illness. The most frequently reported symptoms were headache (60%), rhinorrhea (57%), and myalgias (55%). Gastrointestinal symptoms were less common, with nausea, abdominal pain, and diarrhea reported by only 19%, 21%, and 24% of participants, respectively (Fig 2A). Fever and anosmia were reported in nearly half the participants (48% and 46%, respectively), with fever usually occurring within the first 3-5 days of illness, while anosmia more commonly presenting later in illness, though usually still within the first week. Fifty-eight (40%) participants reported neither fever nor anosmia. Diarrhea, fever, and nausea had the shortest duration of symptoms, with a mean symptom duration of three days or less. Headache, rhinorrhea, and cough had the longest mean duration of around one week (Fig 2B). Examining only household contacts with COVID-19 illness, youth <=21 years (N=33) reported lower cumulative symptoms compared to older adults >30 years (N=44) (median 6 vs 9 symptoms, respectively; p=0.02),

**Fig 2.**
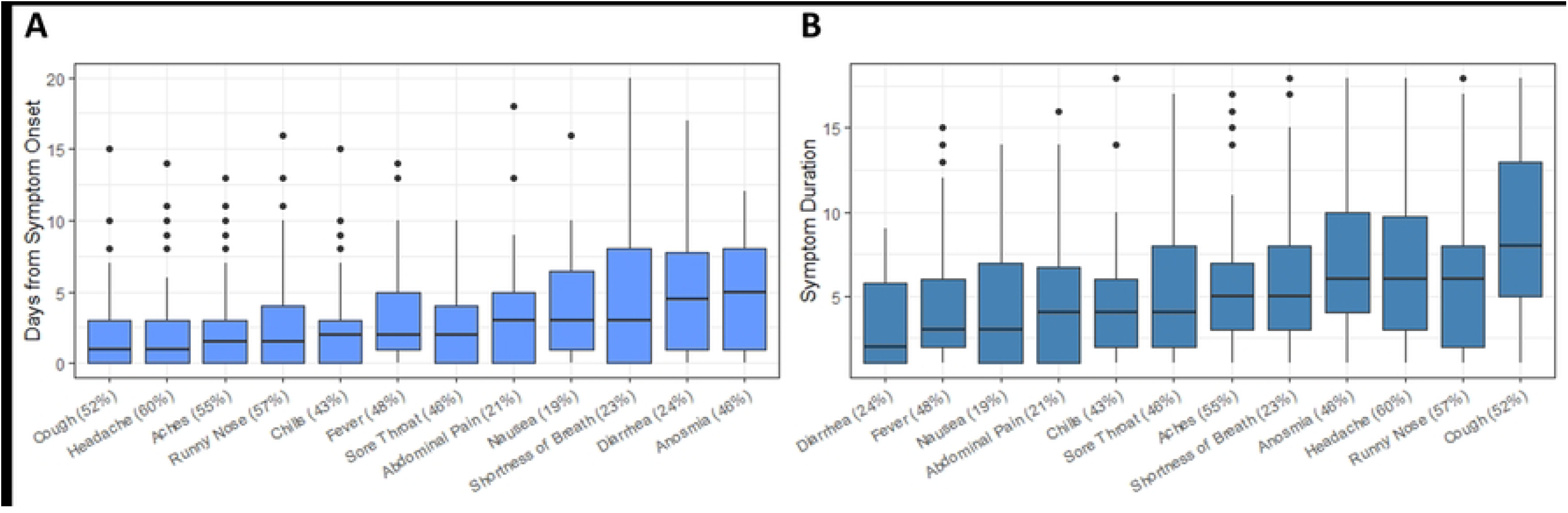
Comparison of symptom frequency, onset, and duration. The frequency of symptoms depicted according to their timing of onset (A) and total duration (B), with median and interquartile ranges.

### Associations with disease severity

Symptom severity increased with age, cardiovascular disease, and viral load (Table 1). Increasing age correlated with increased symptom severity (mean age of 54 years in those who were hospitalized vs. 31 years in those with asymptomatic and mild disease, p=0.003), and the presence of cardiovascular disease (coronary artery disease, hypertension, or diabetes) was associated with severe disease and hospitalization (p=0.02). Higher BMI and male sex seemed to be associated with increased symptom severity, but this finding was not significant in our relatively small sample.

**Table 1:**
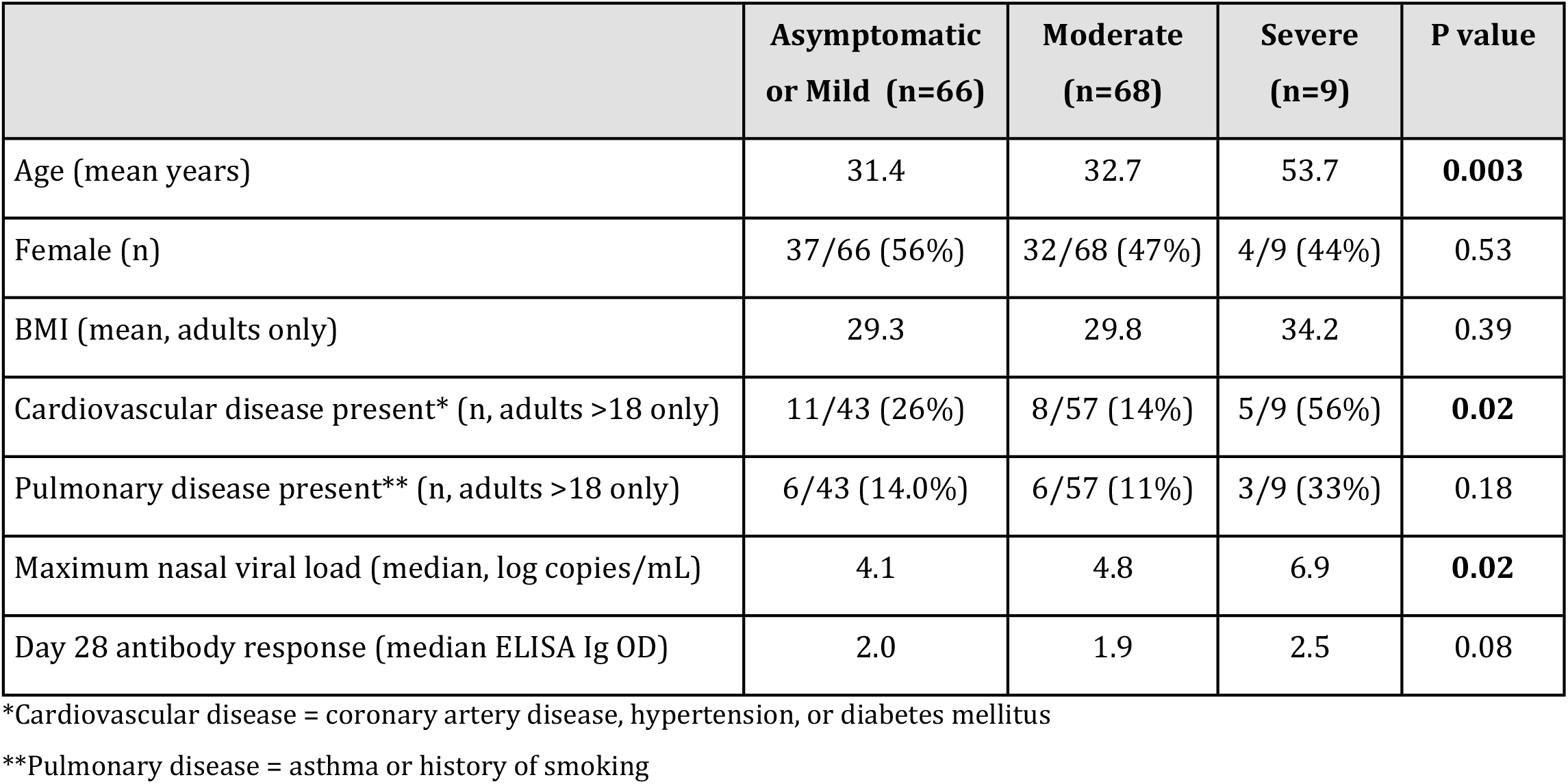
Participant characteristics by symptom severity.

Increased nasal viral load was also associated with increasing symptom severity, with median values of 4.1, 4.8, and 6.9 log copies/mL in asymptomatic/mild, moderate, and severe cases, respectively (Fig S1A). There was a trend towards higher anti-RBD Ig levels at day 28 among those with more severe illness, though this finding did not reach statistical significance (Table 1, Fig S1B).

### Viral load and antibody response

Higher nasal viral load was associated with a higher antibody response in adults, as measured by anti-RBD total Ig levels on day 28 (p<0.001) (Fig S2A). Higher BMI and the presence of cardiovascular disease, defined as report of coronary artery disease, hypertension, or diabetes, among adults were also associated with increased antibody response (p<0.001 for BMI) despite only a weak correlation between BMI and viral load (Fig S2B&C). This association of increased antibody levels in those with higher BMIs persisted even when controlling for symptom severity and the presence of cardiovascular disease (p=0.001). Neither viral load nor antibody response varied by age and sex.

### Symptoms and antibody titers at extended follow-up

Follow-up data was obtained at a single study visit 8-10 months after enrollment, between May 5 and August 27, 2021, for 91 COVID-positive participants out of the 315 individuals enrolled in the original study cohort, including 24 children and 67 adults (Table 2). Of the 91 participants, 45% (41/91) self-rated their COVID-19 illness as having been asymptomatic or mild, compared to 46% of our original cohort that we classified as such. At the time of follow-up, 47/91 (52%) participants were now fully vaccinated.

**Table 2:** Participant characteristics at extended follow-up, occurring a median of 8 months (IQR 8-10) after initial enrollment.

| Participant characteristics | Total N=91<br>Index cases (n=33)<br>Household cases (n=58) |
| --- | --- |
| Male | 38 (42%) |
| Age |  |
| >18 | 67 (74%) |
| 12-18 | 16 (17%) |
| <12 | 8 (8%) |
| Median age (IQR) | 29 (IQR 17-48) |
| Self-rated symptom severity during acute illness |  |
| Asymptomatic | 9 (10%) |
| Mild | 32 (35%) |
| Moderate | 42 (46%) |
| Hospitalized | 3 (3%) |
| Unknown | 5 (5%) |
| Long COVID/PASC (persistent symptoms lasting >6 months) | 15 (16%) |
| Trouble sleeping | 4/15 (27%) |
| Problems with memory | 5/15 (33%) |
| Problems with paying attention | 3/15 (20%) |
| Problems with feeling lightheaded | 2/15 (13%) |
| Periods of racing heart rate | 3/15 (20%) |
| Inability to exercise at pre-COVID level | 4/15 (27%) |
| Inability to return to usual pre-COVID activities | 1/15 (7%) |
| Feeling weak/tired 24-48 hours after activity | 3/15 (20%) |
| Taste and smell not back to normal* | 6/15 (40%) |
| Problems with appetite | 0/15 (0%) |
| Inability to return to work or school | 0/15 (0%) |
| Vaccinated at D180 visit** | 59 (65%) |
| Full | 47/59 (80%) |
| Moderna (partial or full) | 22/59 (37%) |
| Pfizer (partial or full) | 33/59 (56%) |
| Johnson & Johnson (full) | 4/59 (7%) |
| Unvaccinated at D180 visit | 31 (34%) |
| Age <18 | 16/31 (51%) |
| Age 12-18 | 7/31 (23%) |
| Age <12 | 8/31 (26%) |

\*Not specifically elicited but reported as frequent ongoing symptom
\*\*One participant was unaware of their vaccination status because they had participated in a blinded vaccine research study

Fifteen participants (3 children ages 10, 12, and 15; and 12 adults), representing 16% of this follow-up cohort, reported lingering health concerns that they attributed to COVID-19, including the presence of at least one long COVID symptom suggestive of PASC (post-acute sequelae of SARS-CoV-2 infection) that had persisted since the time of their acute infection (Table 2). The most commonly reported ongoing health concerns were taste and smell not back to normal (6 participants), problems with memory (5 participants), trouble sleeping (4 participants), and inability to exercise to pre-COVID levels (4 participants). Additional health issues reported by at least two participants were problems with paying attention, lightheadedness, heart palpitations, feeling weak or tired 24-48 hours after activity, and an alteration of taste or smell.

There was no relationship between the presence of ongoing symptoms suggestive of PASC reported at follow-up and vaccination status, symptom severity during acute infection (n=56), age, sex, BMI, or maximum viral load during acute infection (n=56). However, participants who reported symptoms suggestive of PASC had a higher antibody level on day 28 of their acute infection compared to those who did not report any lingering health concerns related to their COVID illness (median ELISA OD 2.3 (95% CI 2.2-2.6) vs. 1.9 (95% CI 1.3-2.2) in PASC and non-PASC individuals, respectively, p=0.006) (Fig 3). Of those with ELISA OD ≥2.0 at day 28, 26% (9/34) went on to report symptoms of PASC, compared to 7% (3/41) of those with ELISA OD <2.0 at day 28 (p=0.02).

**Fig 3.**
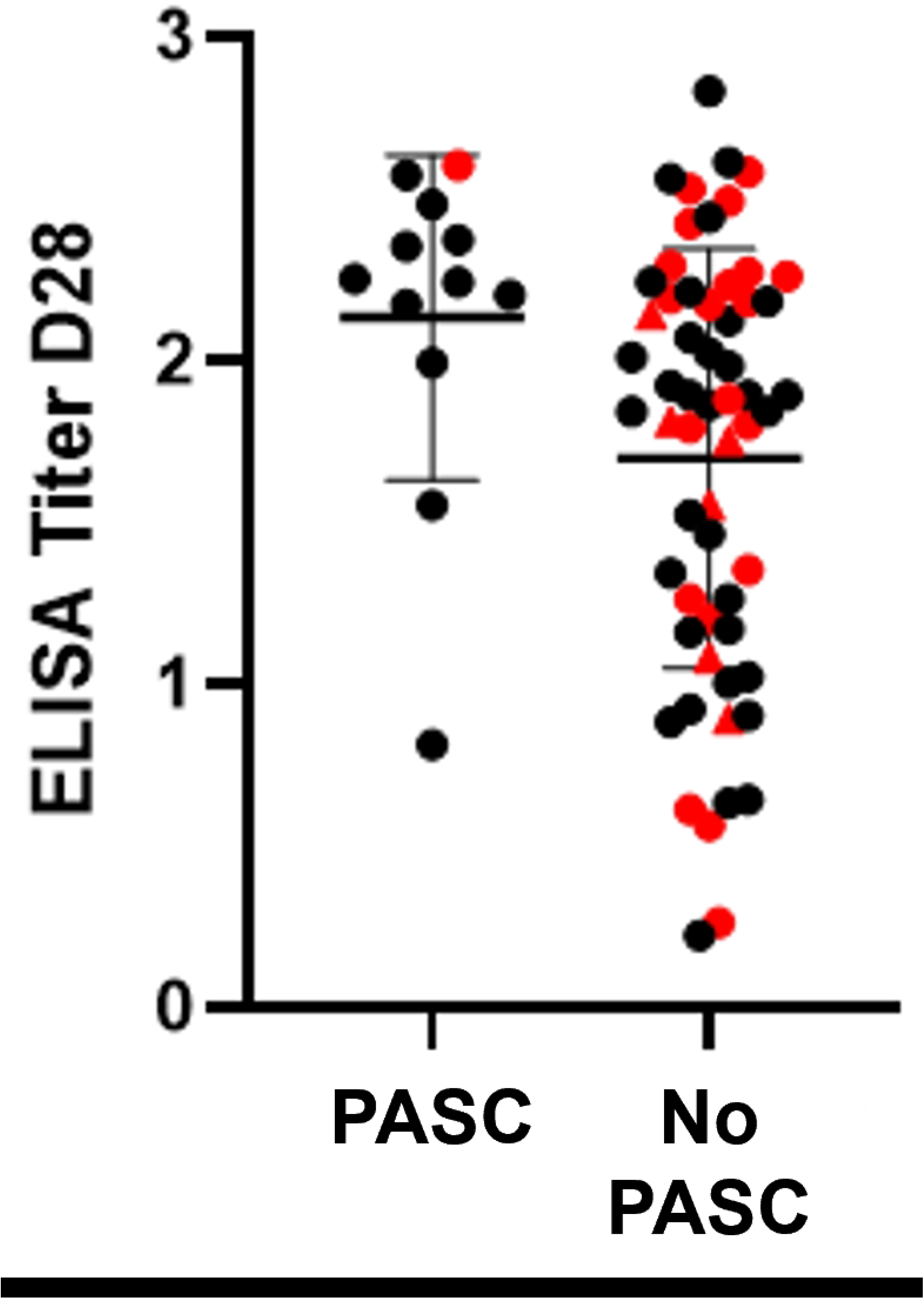
Antibody data at D28 by post-acute sequelae of SARS-CoV-2 (PASC) at 8-10 months. 91 study participants completed an extended follow up visit, of which 15 individuals reported at least one symptom consistent with PASC. Black and red denote vaccinated and unvaccinated people respectively; red triangles are specifically indicate antibody negative at follow-up. The antibody titers at D28 for 12 of these individuals were significantly higher than for individuals with no persisting symptoms (median ELISA OD 2.3 (95% CI 2.2-2.6) vs. 1.91 (95% CI 1.3-2.2) in PASC and non-PASC individuals, respectively, p=0.006). Titers were missing for the other 3 individuals.

As expected, all those who reported a history of vaccination had detectable antibody levels at follow-up. Among 26 participants (9 index cases and 17 household contacts) who remained unvaccinated at the 8-10 month follow-up, 19 (73%) also had detectable antibodies. In this small sample, there was no difference in symptom severity, age, sex, BMI, maximum viral load during acute infection, and antibody response at day 28 between antibody-positive and antibody-negative unvaccinated individuals at the 8-10 month extended visit.

## Discussion

We integrate symptom, viral load, and antibody levels from a sizable and diverse outpatient cohort that underwent daily symptom follow-up during the first six months of the SARS-CoV-2 epidemic in the US, as well as a single long-term follow-up for PASC. Household-based recruitment after enrollment of a symptomatic index case allowed for active surveillance and analysis of SARS-CoV-2 symptoms in 143 participants through the early phase of illness, before the development of antibodies. The bulk of participants (121/143, 85%) had mild or moderate disease. Interestingly, 16% of household contacts were truly asymptomatic, confirmed by daily prospective follow-up through 21 days. This differs from studies on other outpatient cohorts from 2020, which found that most or all patients were pre-symptomatic and not truly asymptomatic [10–12]. This may be attributable to the small size and heterogeneity of populations studied (including residents of long-term skilled nursing facilities) and/or slight differences in symptom ascertainment and scoring, as we still considered participants asymptomatic if they reported one cumulative symptom for the duration of their enrollment, as long the symptom reported was not anosmia, dyspnea, or new or worsening cough.

A study conducted in two hospitals in Hong Kong found that in patients with severe/critical or moderate disease, viral loads in respiratory tract samples peaked in the second week of illness, whereas it peaked in the first week of illness among patients with mild/moderate disease [13]. Our data complements this finding, as patients with more severe disease or requiring hospitalization had symptoms peak around day 9, as opposed to individuals with milder or moderate disease whose symptoms peaked only 3 to 4 days after symptom onset. Time to peak illness of symptoms is not often reported in the research literature but is important for counseling patients. A Canadian study that employed remote monitoring of those quarantining at home found that symptoms peaked 1-2 days later in older people compared to younger persons [14]. Overall, symptoms early in the pandemic were reported to last a median of 2-3 weeks [15,16], though often persisting as long as 3-5 weeks [16–18].

Consistent with other outpatient studies, we found that greater disease severity (more cumulative symptoms) was associated with increased age [10,16,19–21] and the presence of cardiovascular disease (defined as coronary artery disease, hypertension, or diabetes mellitus in our study) [16,21–23]. We also saw nonsignificant trends towards greater disease severity in males and those with higher BMI. Other, but not all, studies have borne out these associations [16,19–22], likely reflecting unmeasured heterogeneity in populations. Overall, the symptom data reported here can serve as a comparative baseline for understanding how symptoms evolved with the evolution of different SARS-CoV-2 variants alongside evolving immunity in the US population.

Unlike most other outpatient cohort studies conducted early in the pandemic in the United States, we integrated viral load and antibody response data alongside information collected from daily symptom diaries. This revealed an association between symptom severity and increased viral loads, supporting a well-established body of evidence from animal models and clinical studies that viral load is a likely determinant of COVID-19 disease severity [23]. Early studies conducted largely in hospitalized patients assessing antibody kinetics showed higher antibody levels in those with more severe disease and higher viral loads [24–27]. In this cohort dominated by non-severe disease, we observed a similar, albeit nonsignificant, trend with increased antibody response with higher viral load in the small group with severe disease (n=9) but no detectable difference between those categorized as having mild or moderate disease. Most, but not all, subsequent studies in diverse community cohorts have confirmed weaker antibody responses in non-hospitalized patients, along with higher neutralizing antibodies in those with severe disease [28–31].

Interestingly, we found that increased BMI was strongly correlated with a higher antibody response in our cohort, seemingly independent of disease severity. Several prior studies have found the same, alongside evidence of over-reactive immune response in obese patients [32–34]. However, there is discordance in the literature since not all could adjust for symptom severity, and some described reduced immune responses in obese patients [35]. In addition to mechanistic studies, further analysis of large non-hospitalized cohorts, also including younger obese patients [36], may shed more light on this question.

Out of 91 COVID-positive patients from the parent study, 15 (16%) individuals noted persistent symptoms attributable to PASC. Like prior data evaluating PASC, many lingering symptoms our patients reported were neurological, such as forgetfulness and sleep disturbances, as well as related to exercise intolerance [37]. Detecting predictive markers for long COVID syndrome has been an important research priority given the millions affected by PASC. In our small sample, we found that a significantly higher proportion of people with ELISA OD ≥2.0 at day 28 reported symptoms of PASC at 8-10 months compared to those with ELISA OD <2.0. This aligns with an association of symptom severity in the acute phase of infection and subsequent PASC described in the STOP-COVID registry, which followed a large outpatient cohort from 2020 in the United States [22]. Large multi-omics and immunologic profiling studies that have sought to find an acute immunological marker of PASC in hospitalized patient populations have highlighted the potential importance of specific autoantibodies in aberrant pathophysiology leading to subsequent long COVID phenotypes [38–40]. Our findings indicate that the potential clinical utility of assessing acute humoral response as a predictor for PASC is not limited to hospitalized patients and should continue to be evaluated in later SARS-CoV-2 variants.

This data was collected before the availability of vaccines or outpatient therapies directed against COVID-19 and thus represents symptoms and disease course in the absence of modifying interventions. It is necessarily limited in its characterization of the earliest D614G SARS-CoV-2 strain that swept the United States in 2020, before the emergence of new COVID-19 variants. Unlike subsequent studies with daily sampling, sampling for nasal viral loads occurred weekly, while antibody responses were measured at study enrollment and 28 days later. Thus the study was limited in its capture of peak measurements and associated viral load and antibody kinetics afforded by more frequent sampling. Although symptom data relied on patient self-report, daily online questionnaires likely minimized recall bias. Regarding detection of PASC, extended follow-up visits were carried out before more formal or comprehensive definitions of PASC were proposed [41]and did not involve in-depth clinical evaluation. Thus, prevalence of PASC may be underestimated or imprecise, limiting investigation of factors associated with PASC. Another limitation is

In conclusion, this study contributes to an understanding of the full spectrum of illness caused by SARS-CoV-2 at different stages in the pandemic, providing early baseline data that can help assess changes in virulence and patterns of disease in the context of evolving variants, stages of vaccination, and increasing availability of treatment for non-hospitalized illness. A significant proportion of participants reported symptoms consistent with PASC, underlying the common occurrence of long COVID symptoms in even mild/moderate illness and the need for further study in these populations. Stored specimens from our study may contribute to such ongoing research [42].

## Data Availability

All relevant data are within the manuscript and its Supporting Information files.

## Acknowledgements

We thank our wonderful CO-HOST study participants, Moby and the Chapel Hill CRS, and the UNC RDC team. Thanks to Michelle Berrey, JoAnn Kuruc, and Dania Munson for help with protocol writing and submission; to Oksana Kharabora, Maureen Furlong, Amy James Loftis, and Dana Swilley for help with study preparation and implementation.

## Supporting Information Captions

**Fig S1. Comparison of symptom severity, viral load, and antibody response**.

**Fig S2. Comparison of antibody response, viral load, and BMI**. Comparison of ELISA OD value on study day 28 and maximum viral load measured by PCR testing (A) and BMI of adults (B) as well as comparison of maximum viral load and BMI of adults (C).

## References

1. To KK-W, Tsang OT-Y, Leung W-S, Tam AR, Wu T-C, Lung DC, et al. Temporal profiles of viral load in posterior oropharyngeal saliva samples and serum antibody responses during infection by SARS-CoV-2: an observational cohort study. Lancet Infect Dis. 2020;20: 565–574.

2. Suh HJ, Kim DH, Heo EY, Lee HW, Lee JK, Lee CS, et al. Clinical characteristics of COVID-19: Clinical dynamics of mild severe acute respiratory syndrome Coronavirus 2 infection detected by early active surveillance. J Korean Med Sci. 2020;35: e297.

3. Sun B, Feng Y, Mo X, Zheng P, Wang Q, Li P, et al. Kinetics of SARS-CoV-2 specific IgM and IgG responses in COVID-19 patients. Emerg Microbes Infect. 2020;9: 940–948.

4. Wu Z, McGoogan JM. Characteristics of and Important Lessons From the Coronavirus Disease 2019 (COVID-19) Outbreak in China: Summary of a Report of 72 314 Cases From the Chinese Center for Disease Control and Prevention. JAMA. 2020;323: 1239–1242.

5. Cerami C, Rapp T, Lin F-C, Tompkins K, Basham C, Muller MS, et al. High household transmission of SARS-CoV-2 in the United States: living density, viral load, and disproportionate impact on communities of color. medRxiv. 2021. doi:10.1101/2021.03.10.21253173

6. Centers for Disease Control and Prevention. CDC 2019-Novel Coronavirus (2019-nCoV) Real-Time RT-PCR Diagnostic Panel. In: CDC 2019-Novel Coronavirus (2019-nCoV) Real-Time RT-PCR Diagnostic Panel [Internet]. Available: https://www.fda.gov/media/134922/download

7. Muller MS, Chhetri SB, Basham C, Rapp T, Lin F-C, Lin K, et al. Practical strategies for SARS-CoV-2 RT-PCR testing in resource-constrained settings. Diagn Microbiol Infect Dis. 2021;101: 115469.

8. Premkumar L, Segovia-Chumbez B, Jadi R, Martinez DR, Raut R, Markmann A, et al. The receptor binding domain of the viral spike protein is an immunodominant and highly specific target of antibodies in SARS-CoV-2 patients. Sci Immunol. 2020;5. doi:10.1126/sciimmunol.abc8413

9. Coronavirus Disease 2019 (COVID-19) 2020 interim case definition, approved august 5, 2020. 15 Aug 2022 [cited 28 Aug 2024]. Available: https://ndc.services.cdc.gov/case-definitions/coronavirus-disease-2019-2020-08-05/

10. Yousaf AR, Duca LM, Chu V, Reses HE, Fajans M, Rabold EM, et al. A prospective cohort study in non-hospitalized household contacts with SARS-CoV-2 infection: symptom profiles and symptom change over time. Clin Infect Dis. 2020. doi:10.1093/cid/ciaa1072

11. Kimball A, Hatfield KM, Arons M, James A, Taylor J, Spicer K, et al. Asymptomatic and Presymptomatic SARS-CoV-2 Infections in Residents of a Long-Term Care Skilled Nursing Facility - King County, Washington, March 2020. MMWR Morb Mortal Wkly Rep. 2020;69: 377–381.

12. Arons MM, Hatfield KM, Reddy SC, Kimball A, James A, Jacobs JR, et al. Presymptomatic SARS-CoV-2 Infections and Transmission in a Skilled Nursing Facility. N Engl J Med. 2020;382: 2081–2090.

13. Lui G, Ling L, Lai CK, Tso EY, Fung KS, Chan V, et al. Viral dynamics of SARS-CoV-2 across a spectrum of disease severity in COVID-19. J Infect. 2020;81: 318–356.

14. Gershon AS, Patel N, Liaqat S, Liaqat D, de Lara E, To T, et al. Symptom Burden in Patients of Different Ages with Acute COVID-19 Infection Quarantining at Home. B57 CUTTING EDGE COVID RESEARCH. American Thoracic Society; 2022. pp. A3170–A3170.

15. Lane A, Hunter K, Lee EL, Hyman D, Bross P, Alabd A, et al. Clinical characteristics and symptom duration among outpatients with COVID-19. Am J Infect Control. 2022;50: 383–389.

16. Blair PW, Brown DM, Jang M, Antar AAR, Keruly JC, Bachu VS, et al. The Clinical Course of COVID-19 in the Outpatient Setting: A Prospective Cohort Study. Open Forum Infect Dis. 2021;8: ofab007.

17. Cellai M, O’Keefe JB. Characterization of Prolonged COVID-19 Symptoms in an Outpatient Telemedicine Clinic. Open Forum Infect Dis. 2020;7: ofaa420.

18. Tenforde MW, Billig Rose E, Lindsell CJ, Shapiro NI, Files DC, Gibbs KW, et al. Characteristics of Adult Outpatients and Inpatients with COVID-19 - 11 Academic Medical Centers, United States, March-May 2020. MMWR Morb Mortal Wkly Rep. 2020;69: 841–846.

19. Nguyen NT, Chinn J, Kirby K, Hohmann SF, Amin A. Outcomes of COVID-19 adults managed in an outpatient versus hospital setting. PLoS One. 2022;17: e0263813.

20. Pettrone K, Burnett E, Link-Gelles R, Haight SC, Schrodt C, England L, et al. Characteristics and Risk Factors of Hospitalized and Nonhospitalized COVID-19 Patients, Atlanta, Georgia, USA, March-April 2020. Emerg Infect Dis. 2021;27: 1164–1168.

21. Halalau A, Odish F, Imam Z, Sharrak A, Brickner E, Lee PB, et al. Epidemiology, Clinical Characteristics, and Outcomes of a Large Cohort of COVID-19 Outpatients in Michigan. Int J Gen Med. 2021;14: 1555–1563.

22. Pływaczewska-Jakubowska M, Chudzik M, Babicki M, Kapusta J, Jankowski P. Lifestyle, course of COVID-19, and risk of Long-COVID in non-hospitalized patients. Front Med. 2022;9: 1036556.

23. da Silva SJR, de Lima SC, da Silva RC, Kohl A, Pena L. Viral Load in COVID-19 Patients: Implications for Prognosis and Vaccine Efficacy in the Context of Emerging SARS-CoV-2 Variants. Front Med. 2021;8: 836826.

24. Zhang X, Lu S, Li H, Wang Y, Lu Z, Liu Z, et al. Viral and antibody kinetics of COVID-19 patients with different disease severities in acute and convalescent phases: A 6-month follow-up study. Virol Sin. 2020;35: 820–829.

25. Masiá M, Telenti G, Fernández M, García JA, Agulló V, Padilla S, et al. SARS-CoV-2 Seroconversion and Viral Clearance in Patients Hospitalized With COVID-19: Viral Load Predicts Antibody Response. Open Forum Infect Dis. 2021;8: ofab005.

26. Wang Y, Zhang L, Sang L, Ye F, Ruan S, Zhong B, et al. Kinetics of viral load and antibody response in relation to COVID-19 severity. J Clin Invest. 2020;130: 5235–5244.

27. Li L, Liang Y, Hu F, Yan H, Li Y, Xie Z, et al. Molecular and serological characterization of SARS-CoV-2 infection among COVID-19 patients. Virology. 2020;551: 26–35.

28. Sancilio A, Schrock JM, Demonbreun AR, D’Aquila RT, Mustanski B, Vaught LA, et al. COVID-19 symptom severity predicts neutralizing antibody activity in a community-based serological study. Sci Rep. 2022;12: 12269.

29. Horton DB, Barrett ES, Roy J, Gennaro ML, Andrews T, Greenberg P, et al. Determinants and Dynamics of SARS-CoV-2 Infection in a Diverse Population: 6-Month Evaluation of a Prospective Cohort Study. J Infect Dis. 2021;224: 1345–1356.

30. Imai K, Kitagawa Y, Tabata S, Kubota K, Nagura-Ikeda M, Matsuoka M, et al. Antibody response patterns in COVID-19 patients with different levels of disease severity in Japan. J Med Virol. 2021;93: 3211–3218.

31. Hung IF-N, Cheng VC-C, Li X, Tam AR, Hung DL-L, Chiu KH-Y, et al. SARS-CoV-2 shedding and seroconversion among passengers quarantined after disembarking a cruise ship: a case series. Lancet Infect Dis. 2020;20: 1051–1060.

32. Onyango TB, Zhou F, Bredholt G, Brokstad KA, Lartey S, Mohn KG-I, et al. SARS-CoV-2 specific immune responses in overweight and obese COVID-19 patients. Front Immunol. 2023;14: 1287388.

33. Zhai B, Clarke K, Bauer DL, Moehling Geffel KK, Kupul S, Schratz LJ, et al. SARS-CoV-2 Antibody Response Is Associated with Age and Body Mass Index in Convalescent Outpatients. J Immunol. 2022;208: 1711–1718.

34. Soffer S, Glicksberg BS, Zimlichman E, Efros O, Levin MA, Freeman R, et al. The Association Between Obesity and Peak Antibody Titer Response in COVID-19 Infection. Obesity. 2021. doi:10.1002/oby.23208

35. Tong MZ, Sng JD, Carney M, Cooper L, Brown S, Lineburg KE, et al. Elevated BMI reduces the humoral response to SARS-CoV-2 infection. Clin Transl Immunology. 2023;12: e1476.

36. Nilles EJ, Siddiqui SM, Fischinger S, Bartsch YC, de St Aubin M, Zhou G, et al. Epidemiological and Immunological Features of Obesity and SARS-CoV-2. Viruses. 2021;13. doi:10.3390/v13112235

37. Robineau O, Zins M, Touvier M, Wiernik E, Lemogne C, de Lamballerie X, et al. Long-lasting symptoms after an acute COVID-19 infection and factors associated with their resolution. JAMA Netw Open. 2022;5: e2240985.

38. Su Y, Yuan D, Chen DG, Ng RH, Wang K, Choi J, et al. Multiple early factors anticipate post-acute COVID-19 sequelae. Cell. 2022;185: 881–895.e20.

39. Ozonoff A, Jayavelu ND, Liu S, Melamed E, Milliren CE, Qi J, et al. Features of acute COVID-19 associated with post-acute sequelae of SARS-CoV-2 phenotypes: results from the IMPACC study. Nat Commun. 2024;15: 216.

40. Leung JM, Wu MJ, Kheradpour P, Chen C, Drake KA, Tong G, et al. Early immune factors associated with the development of post-acute sequelae of SARS-CoV-2 infection in hospitalized and non-hospitalized individuals. Front Immunol. 2024;15: 1348041.

41. Ely EW, Brown LM, Fineberg HV, National Academies of Sciences, Engineering, and Medicine Committee on Examining the Working Definition for Long Covid. Long covid defined. N Engl J Med. 2024 [cited 29 Sep 2024]. doi:10.1056/NEJMsb2408466

42. Figueiredo JC, Hirsch FR, Kushi LH, Nembhard WN, Crawford JM, Mantis N, et al. Mission, Organization, and Future Direction of the Serological Sciences Network for COVID-19 (SeroNet) Epidemiologic Cohort Studies. Open Forum Infect Dis. 2022;9: ofac171.

